# Effectiveness of vaccination against SARS-CoV-2 infection and Covid-19 hospitalization among Finnish elderly and chronically ill – An interim analysis of a nationwide cohort study

**DOI:** 10.1101/2021.06.21.21258686

**Authors:** Ulrike Baum, Eero Poukka, Arto A. Palmu, Heini Salo, Toni O. Lehtonen, Tuija Leino

## Abstract

**Background:** In Finland, both mRNA and adenovirus vector (AdV) Covid-19 vaccines have been used after the vaccination campaign started on December 27, 2020. Vaccination of the elderly and chronically ill was prioritized and the interval between doses set to 12 weeks. The objective of this interim analysis was to evaluate first and second dose vaccine effectiveness (VE) in a real-world setting.

**Methods:** During the first 5 months of the campaign, a register-based cohort study was conducted in the Finnish elderly aged 70+ years and those aged 16–69 years with medical conditions predisposing to severe Covid-19 (chronically ill). Using Cox regression, VE against SARS-CoV-2 infection and Covid-19 hospitalization was estimated comparing the hazard in the vaccinated with that in the unvaccinated.

**Results:** The cohorts included 901092 elderly (89% vaccinated) and 774526 chronically ill (69% vaccinated) individuals. Three weeks after the first dose, mRNA VE against infection was 45% (95% confidence interval, 36–53%) and 40% (26–51%) in elderly and chronically ill; mRNA VE against hospitalization was 63% (49–74%) and 82% (56–93%). In chronically ill, AdV VE was 42% (32–50) and 62% (42– 75%) against infection and hospitalization, respectively. One week after the second dose, mRNA VE against infection was 75% (65–82%) and 77% (65–85%) in elderly and chronically ill; mRNA VE against hospitalization was 93% (70–98%) and 90% (29–99%).

**Conclusions:** Covid-19 vaccines protect against SARS-CoV-2 infection and Covid-19 hospitalization. A single dose provides moderate protection in elderly and chronically ill, although 2 doses are clearly superior.

**summary:** This register-based cohort study demonstrates that Covid-19 vaccines protect against SARS-CoV-2 infection and Covid-19 hospitalization among Finnish elderly and chronically ill. Vaccine effectiveness against hospitalization was moderate after 1 dose and increased to ≥90% after 2 doses.

## Introduction

The global burden of the ongoing Covid-19 pandemic caused by the severe acute respiratory syndrome coronavirus 2 (SARS-CoV-2) has been immense. Finland, a Nordic country with 5.5 million inhabitants, has been less affected than the rest of Europe [1]. As of May 24, 2021, there have been roughly 90 000 confirmed Covid-19 cases and 942 Covid-19-associated deaths in the Finnish population [2].

In late 2020 the European Medicines Agency authorized the first Covid-19 vaccines for use in the European Union [3]. Finland started its vaccination campaign promptly after, on December 27, prioritizing healthcare workers treating Covid-19 patients as well as residents and healthcare workers of long-term care facilities [4]. Thereafter, the elderly aged 70+ years and individuals aged 16+ years with (highly) predisposing medical conditions for severe Covid-19 were vaccinated [4]. Due to vaccine shortage, Finland decided to postpone the second dose until 12 weeks after the first, similarly as in the United Kingdom (UK) [5], to provide more people with at least 1 dose as early as possible. Therefore, most of the elderly and chronically ill were vaccinated only once in the first quarter of 2021 [6]. By May 24, 2 messenger ribonucleic acid (mRNA) vaccines (by BioNTech/Pfizer and Moderna) and 2 adenovirus vector (AdV) vaccines (by Oxford/AstraZeneca and Janssen) have been approved by the European Medicines Agency [7] but only the first 3 have so far been used in Finland.

The first cohort studies assessing the effectiveness of Covid-19 vaccines were published in early 2021. The vaccine effectiveness (VE) estimates for 2 doses from BioNTech/Pfizer varied between 86% in healthcare workers and 92% in nationwide settings and, thus, were close to the vaccine efficacy measured in randomized controlled trials [8,9]. However, VE in the elderly and immunocompromised individuals has been suspected to be lower [10].

The aim of this observational study was to evaluate the interim effectiveness of Covid-19 vaccines in 2 cohorts, i.e. the elderly and the chronically ill, during the first 5 months of the vaccination campaign in Finland. As the administration of the second dose has been postponed, we focus especially on how the VE is sustained after the first dose.

## Methods

To estimate effectiveness of Covid-19 vaccines in the elderly and the chronically ill in Finland, we conducted a cohort study based on data from 8 national registers linked using the unique Finnish personal identity code. The study follow-up begun with the first Covid-19 vaccinations in Finland (December 27, 2020) and ended on May 24, 2021. The 2 population-based study cohorts were the Finnish elderly aged 70+ years and the chronically ill aged 16–69 years at the beginning of the study. All study subjects were permanent residents and registered in the Population Information System, which provided information on each individual’s date of birth and death (if applicable), sex and municipality of residence.

The cohort of the chronically ill was formed by individuals identified to be at increased risk of severe Covid-19 due to certain medical conditions. These conditions included active cancer treatment, organ or stem cell transplant, severe disorders of the immune system, severe chronic renal, liver or pulmonary disease, type 1 and type 2 diabetes, Down syndrome, severe heart disease, neurological illness or condition affecting breathing, immunosuppressive drug therapy for autoimmune disease, adrenal gland disorders, sleep apnea and psychotic disorders (Supplementary Table 1).

Distinguishing between predisposing and highly predisposing conditions (Supplementary Table 1), presence of the aforementioned medical conditions was determined in both cohorts using the diagnostic data in the Care Register for Health Care and the Register of Primary Health Care Visits since 2015 as well as the data in the Special Reimbursement Register and Prescription Centre database maintained by the Social Insurance Institution. The Special Reimbursement Register allowed the identification of all individuals entitled, at least until January 1, 2020, to a special reimbursement for medical expenses, indicating that the entitled individual met explicit criteria for selected medical conditions (Supplementary Table 1). The Prescription Centre database was used to identify all individuals using selected medications of interest (Supplementary Table 1).

The outcomes of interest were SARS-CoV-2 infection, laboratory-confirmed through polymerase chain reaction or antigen test as registered in the National Infectious Diseases Register, and Covid-19 hospitalization, recorded in the Care Register for Health Care, timely associated with a confirmed SARS-CoV-2 infection. Covid-19 hospitalization was defined as any inpatient hospital admission with Covid-19 (International Classification of Diseases, tenth revision, codes: U07.1, U07.2), acute respiratory tract infections (J00–J22, J46) or severe complications of lower respiratory tract infections (J80–84, J85.1, J86) as primary or secondary diagnosis. Hospital admissions and SARS-CoV-2 infections were timely associated if the infection was confirmed up to 14 days before or 7 days after the admission. Individuals with a confirmed SARS-CoV-2 infection prior to the study follow-up were excluded from the cohorts.

The exposure of interest was vaccination against Covid-19 with at least 1 dose of any vaccine, exactly 1 or 2 doses of mRNA vaccine, or exactly 1 or 2 doses of AdV vaccine (chronically ill only). The vaccine and date of vaccination were extracted from the National Vaccination Register. The time since vaccination with the first dose was split into the intervals 0–6 days since vaccination (DSV), 7–13 DSV, 14–20 DSV, 21–27 DSV, 28–34 DSV, 35–41 DSV and 42+ DSV. The time since vaccination with the second dose was split into the intervals 0–6 DSV and 7+ DSV.

Each study subject was considered to be at risk of the outcome of interest (confirmed SARS-CoV-2 infection or Covid-19 hospitalization) from the beginning of the study until the first occurrence of any of the following events: outcome of interest, death, end of study, or day 14 after confirmed SARS-CoV-2 infection (if the outcome of interest was Covid-19 hospitalization). Using Cox regression with time in study as the underlying time scale, we compared the hazard of confirmed SARS-CoV-2 infection or Covid-19 hospitalization in vaccinated study subjects with the hazard in the unvaccinated. The effect measure of interest was VE, quantified as 1 minus the hazard ratio adjusted for age group (chronically ill: 16–38, 39–51, 52–59, 60–64, 65–69 years (age distribution quintiles); elderly: 70–74, 75–79, 80–89, 90+ years), sex, presence of medical conditions predisposing to severe Covid-19 (Supplementary Tables 1, 2 and 3), residence in the most affected hospital district Helsinki-Uusimaa (1.7 million inhabitants), and residence in long-term care (elderly only).

All analyses were performed in R 3.6.3 (R Foundation for Statistical Computing, Vienna, Austria).

## Results

The study cohorts of the elderly and the chronically ill included 901 092 and 774 526 individuals, respectively, of which 89% and 69% were vaccinated at least once against Covid-19 during the study follow-up. More than 90% of the vaccinated elderly had received mRNA vaccine, while among the chronically ill 2 thirds of the vaccinees had received mRNA vaccine; the rest had received AdV vaccine from Oxford/AstraZeneca (Figure 1). Only a small proportion was vaccinated twice (Figure 1). In the elderly, the vaccination coverage was similar across age groups, sex, and hospital districts (Supplementary Table 2). In the chronically ill, the vaccination coverage increased with age (Supplementary Table 3).

**Figure 1:**
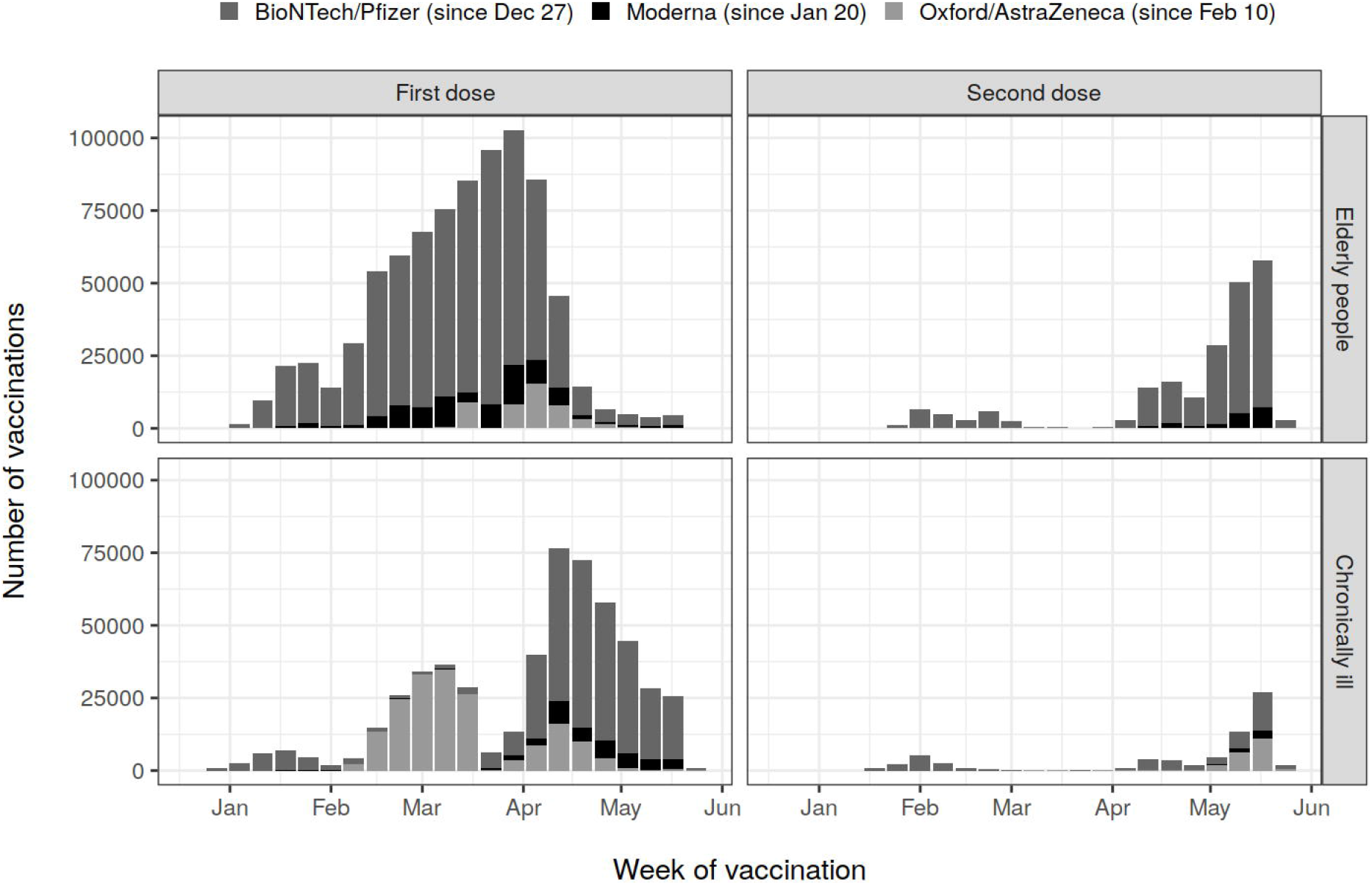
Number of vaccinations administered in the 2 study cohorts of Finnish elderly aged 70+ years and chronically ill aged 16–69 years by vaccine manufacturer and week of vaccination.

In total, there were 2265 cases of confirmed SARS-CoV-2 infection and 466 cases of Covid-19 hospitalization among the elderly and 5929 cases of confirmed infection and 598 cases of hospitalization among the chronically ill. The VE against confirmed infection 21+ days since the first vaccination with any vaccine was 51% (95% confidence interval: 44–58%) in the elderly and 48% (41–54%) in the chronically ill. The corresponding VE against hospitalization was 66% (53– 76%) and 69% (55–79%), respectively.

In both cohorts, the hazards of confirmed infection and hospitalization during the follow-up of less than 7 days since first vaccination were significantly lower than the corresponding hazards in the unvaccinated follow-up (Supplementary Tables 4, 5 and 6). This initial difference between the vaccinated and unvaccinated decreased during the first 2–3 weeks. VE estimates against confirmed infection and hospitalization started to rise 14 and 21 days after vaccination.

In the elderly, the effectiveness of exactly 1 dose of any mRNA vaccine against confirmed infection and hospitalization 21+ DSV was 45% (95% confidence interval: 36–53%) and 63% (49–74%) respectively. After the first three weeks since vaccination, VE against infection was constant, while VE against hospitalization further increased over time (Table 1). After the second mRNA vaccination, VE against confirmed infection and hospitalization was 75% (65–82%) and 93% (70– 98%) (Table 1).

**Table 1:**
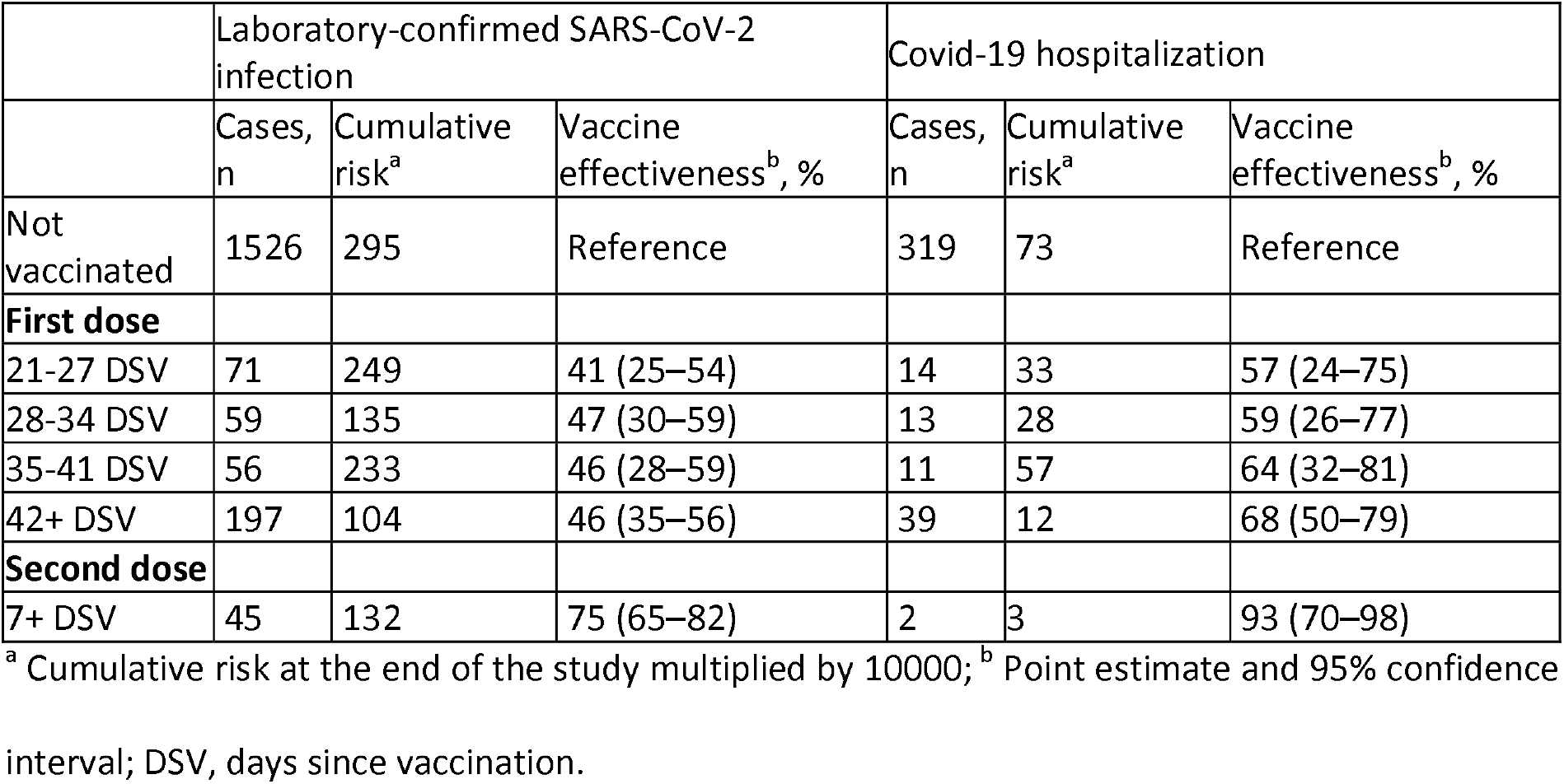
mRNA vaccine effectiveness in the Finnish elderly aged 70+ years adjusted for age group, sex, presence of medical conditions, residence in Helsinki-Uusimaa hospital district and residence in long-term care.

In the chronically ill, the effectiveness of exactly 1 dose of any mRNA vaccine against confirmed infection and hospitalization 21+ DSV was 40% (95% confidence interval: 26–51%) and 82% (56– 93%), respectively. Although initially constant, VE appeared to decrease after 42 days (Table 2). However, the confidence intervals are broad and overlapping. After the second mRNA vaccination, VE against confirmed infection and hospitalization was 77% (65–85%) and 90% (29–99%) (Table 2).

**Table 2:** mRNA vaccine effectiveness in the chronically ill aged 16–69 years adjusted for age group, sex, presence of medical conditions and residence in Helsinki-Uusimaa hospital district.

|  | Laboratory-confirmed SARS-CoV-2 infection |  |  | Covid-19 hospitalization |  |  |
| --- | --- | --- | --- | --- | --- | --- |
|  | Cases, n | Cumulative risk<sup>a</sup> | Vaccine effectiveness<sup>b</sup>, % | Cases, n | Cumulative risk<sup>a</sup> | Vaccine effectiveness<sup>b</sup>, % |
| Not vaccinated | 5280 | 859 | Reference | 528 | 90 | Reference |
| **First dose** |  |  |  |  |  |  |
| 21-27 DSV | 35 | 242 | 41 (17–58) | 1 | 2 | 89 (24–99) |
| 28-34 DSV | 20 | 217 | 58 (34–73) | 1 | 2 | 86 (-1 to 98) |
| 35-41 DSV | 13 | 294 | 59 (29–76) | 0 | 0 | Not estimated |
| 42+ DSV | 43 | 431 | 14 (-16 to 37) | 3 | 13 | 59 (-29 to 87) |
| **Second dose** |  |  |  |  |  |  |
| 7+ DSV | 20 | 181 | 77 (65–85) | 1 | 32 | 90 (29–99) |

<sup>a</sup> Cumulative risk at the end of the study multiplied by 10000; <sup>b</sup> Point estimate and 95% confidence interval; DSV, days since vaccination.

The effectiveness of exactly 1 dose of AdV vaccine against confirmed infection and hospitalization 21+ DSV was 42% (95% confidence interval: 32–50%) and 62% (42–75%), respectively. The VE estimates fluctuated over time but did not decrease after 42 days (Table 3). Due to the small number of vaccinees given 2 doses of AdV vaccine (Figure 1), the effectiveness of the second AdV vaccination could not be estimated (Table 3).

**Table 3:**
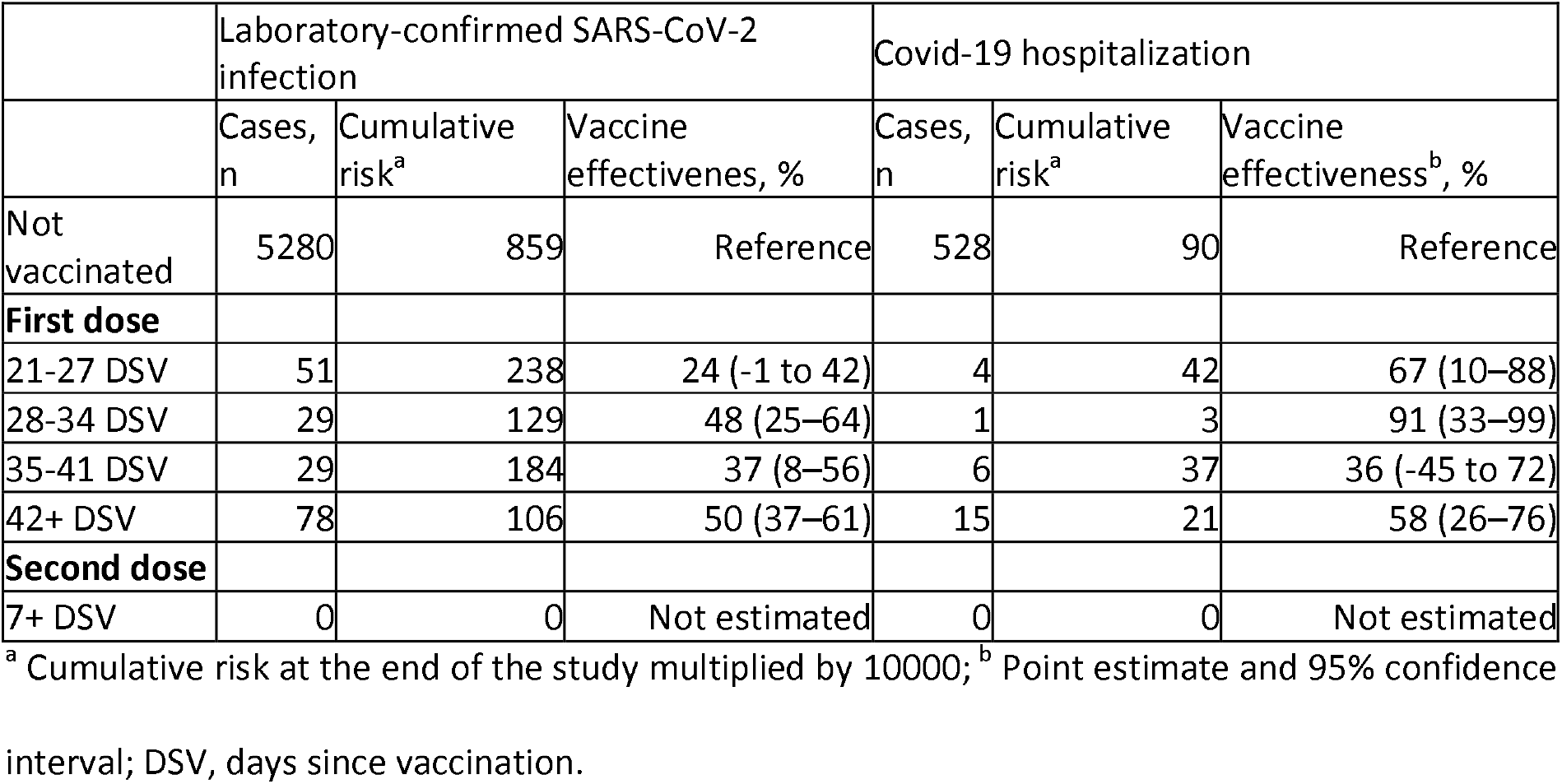
Adenovirus vector vaccine effectiveness in the chronically ill aged 16–69 years adjusted for age group, sex, presence of medical conditions and residence in Helsinki-Uusimaa.

## Discussion

The present observational, population-based study quantified the interim effectiveness of Covid-19 vaccines in the elderly and the chronically ill during the first 5 months of the vaccination campaign in Finland. Three weeks after the first dose, VE was moderate against confirmed SARS-CoV-2 infection but considerably higher against Covid-19 hospitalization. After the first mRNA vaccination (BioNTech/Pfizer or Moderna), the VE estimates were constant in the elderly from the fourth through the seventh week and beyond. However, we were not able to demonstrate sustained mRNA VE in the chronically ill for all post-vaccination intervals. Nevertheless, the second mRNA vaccination increased the VE estimates substantially, both in the elderly and the chronically ill. After the first AdV vaccination (Oxford/AstraZeneca), the observed VE sustained through all post-vaccination intervals in the study but VE of the second dose could not be estimated.

Several studies have previously evaluated VE against SARS-CoV-2 infection. An observational, population-based study from Israel estimated VE among the elderly aged 70+ years at 50 and 95% 3 weeks after the first dose and 1 week after the second dose of the BioNTech/Pfizer vaccine, respectively [8]. Although similar, these results might, however, not be fully comparable to our results as all individuals confined to their homes or long-term care facilities were excluded from the Israeli study but included in ours. In another study from Israel, VE against asymptomatic and symptomatic infection was 52% and 63%, respectively, in the third week after the first dose [11]. In England, VE against symptomatic infection in elderly aged 70+ years was estimated at 50% in the fifth week after the first dose from BioNTech/Pfizer and 61% after the first dose from Oxford/AstraZeneca [12]. In agreement with our findings, the results by Lopez Bernal et al. [12] thus indicate that VE levels sustain even beyond the first 3 weeks after the first dose.

Previous studies have also assessed VE against more severe outcomes and, in line with our study, detected higher protection levels against Covid-19 hospitalization than infection. In the third week after the first dose from BioNTech/Pfizer, VE against hospitalization was found to be 74% and 76% in the general Israeli population aged 16+ years [8,11]. In the subsequent weeks VE of the first BioNTech/Pfizer dose ranged between 77 and 91% according to a Scottish cohort study [13]. When stratified by age, the VE after 7 weeks was 76% among those aged 65–79 years and 85% among those aged 80+ years [13]. In addition, VE was estimated at 77% 2 weeks after the first mRNA vaccination in a population with history of prior all-cause hospitalization in the United States [14]. While the results of those studies are similar, our findings suggest that the VE of only one dose might be lower. Nevertheless, the present estimates concerning the VE of two doses are comparable to the previously published figures ranging between 87 and 97% [8,11,14,15].

Although a direct comparison of the effectiveness estimates for mRNA and AdV vaccines may be influenced by the different dates the vaccines were introduced in the population (Figure 1), we did not detect meaningful differences. This finding is in line with other studies that analyzed the effectiveness of both mRNA and AdV vaccines against SARS-CoV-2 infection [12] and Covid-19 hospitalization [12,13,16].

Although our VE estimates were roughly similar to those obtained in previous studies [8,11-14], our estimates appear to be at the lower side of the spectrum. One reason might be that in Finland vaccinations were given first to the oldest among the elderly and the most immunocompromised among the chronically ill. High age seems to affect the effectiveness of Covid-19 vaccines less than expected based on the experience with other vaccines [8,10,11,13]. Nevertheless, VE was found to be lower among the very old, the immunocompromised and residents of long-term care facilities [10,17] and to vary across different predisposing medical conditions [10,18].

In Finland, Alpha was the predominant SARS-CoV-2 variant causing 60–70% of all SARS-CoV-2 infections in the capital region and 50% of all SARS-CoV-2 infections in the rest of the country in March 2021. At the same time, Beta caused only 5% of the infections [19], but local reports indicate that the incidence of Beta has increased. Recently, there has been a concern that certain SARS-CoV-2 variants are associated with higher transmissibility [20], mortality [21], antibody resistance [22] and capability to cause Covid-19 epidemics in a population with high seroprevalence [23]. It has been shown that Covid-19 vaccines are effective against Alpha [11,24,25] but their efficacy against Beta seems to be reduced [26]. VE in Finland might, therefore, be decreased due to the increased incidence of Beta.

The main strengths of this analysis are the size and representativeness of the 2 cohorts and the register-based nature of the study. In Finland, the recording of Covid-19 vaccinations into the patient information system is mandatory, as is the notification of SARS-CoV-2 positive laboratory findings. These data are then automatically transferred into the national registers. Because exposure, outcome and covariate data are recorded directly at the source and independently, the chance of measurement and recall bias has been minimized. By using Cox regression with time in study as the underlying time scale, we controlled any temporal changes in the hazard of infection and disease or detection of those. Moreover, it can be assumed that a great proportion of symptomatic infections have been captured since the threshold for testing was very low and testing capacities were at no point exhausted. Covid-19 testing has been free of charge and its importance emphasized through official channels.

A limitation of this study is the unknown proportion of asymptomatic cases among the confirmed infections. As VE is greater when measured against a more severe outcome [8], the proportion of asymptomatic cases strongly influences the VE estimates. The major weakness is, however, the presence of time-limited selection bias as in [13]. At the day of vaccination, vaccinees were healthier than average due to the policy to postpone the vaccination of individuals with acute symptoms suggestive of Covid-19 and those in self-isolation. Consequently, the hazards of infection and hospitalization in the recently vaccinated were lower than the hazards in the unvaccinated. In the absence of any bias, these hazards should have been similar in the vaccinated and unvaccinated as it takes 2–3 weeks for the vaccine-induced immune response to develop. In addition, residual confounding may be present due to the observational nature of the study.

## Conclusions

Covid-19 vaccines protect against SARS-CoV-2 infection and Covid-19 hospitalization. We detected no difference between mRNA and AdV vaccines. In the elderly and the chronically ill, a single dose provides moderate protection even beyond the first 3 weeks since vaccination, yet the full series of 2 doses is superior. Further studies are needed to assess Covid-19 vaccine effectiveness for different brands, schedules and target groups.

## Supporting information

Ethical concerns

## Data Availability

By Finnish law, the authors are not permitted to share individual-level register data. The computing code is available upon request.

## Acknowledgements

The authors thank Esa Ruokokoski, Jonas Sundman, Oskari Luomala, Teemu Möttönen and Tuomo Nieminen from the Finnish Institute for Health and Welfare for their assistance in managing the study data.

## Authors’ contributions

UB, TL and AAP conceptualized the study. HS and TOL identified the medical conditions predisposing to severe Covid-19. UB conducted the statistical analysis. EP and TL reviewed the literature. UB and EP drafted the manuscript.

