## Supplementary tables 1-6 for "Effectiveness of vaccination against SARS-CoV-2 infection and Covid-19 hospitalization among Finnish elderly and chronically ill – An interim analysis of a nationwide cohort study"

**Supplementary Table 1:** Definition of medical conditions (highly) predisposing to severe Covid-19.

| Medical condition | Classification | Codes | Register |
|---|---|---|---|
| **Highly predisposing to severe Covid-19** | | | |
| Organ or stem cell transplant | ICD-10 | T86, Z94 | 1,2 |
| Active cancer treatment | ICD-10 | C00–C97 (except C44), D05.1, D39 | 1,2 |
| Severe disorders of the immune system | ICD-10 | D70.8, D80–D84, E31.00 | 2 |
| Severe chronic renal disease | ICD-10 | I12, I13, N00–N05, N07, N08, N11, N14, N18, N19, E10.2, E11.2, E14.2 | 1,2 |
| Asthma requiring continuous medication | ICD-10 | J45, J46 | 2, 3 |
| | ICPC-2 | R96 | 3 |
| Severe chronic pulmonary disease | ICD-10 | J41–J44, J47, Z90.2 | 2 |
| Type 2 diabetes requiring medication | ICD-10 | E11, E13, E14 | 2,3 |
| | ICPC-2 | T90 | 3 |
| Blood glucose lowering drugs, excluding insulin | ATC | A10B | 4 |
| Down syndrome | ICD-10 | Q90 | 2, 3 |
| **Predisposing to severe Covid-19** | | | |
| Severe heart disease | ICD-10 | I11–I13, I15, I20–I25, I50 | 1, 2 |
| Neurological illness or condition that affects breathing | ICD-10 | G70-G73, G80–G83, I60–I69 | 2 |
| Immunosuppressive drug therapy for autoimmune disease | | | |
| Autoimmune disease | ICD-10 | D86, K50, K51, L40, M02, M05–M07, M13.9, M45, M46.0, M46.1, M46.9, M94.1 | 1, 2 |
| Immunosuppressive drug therapy | ATC | H02AB02, H02AB04, H02AB06, H02AB07, L01BA01, L01XC02, L04AA06, L04AA10, L04AA13, L04AA18, L04AA24, L04AA26, L04AA29, L04AA33, L04AA37, L04AB, L04AC, L04AD01, L04AD02, L04AX01, L04AX03 | 4 |
| Severe chronic liver disease | ICD-10 | K70.2, K70.3, K70.4, K71–K74 | 2 |
| Type 1 diabetes | ICD-10 | E10 | 2, 3 |
| | ICPC-2 | T89 | 3 |
| Insulin and analogues | ATC | A10A | 4 |
| Adrenal insufficiency | ICD-10 | E25.0, E27.1, E27.2, E27.4, E31.00, E31.01, E31.08, E89.6 | 1, 2 |
| Sleep apnea | ICD-10 | G47.3 | 2, 3 |
| Continuous positive airway pressure therapy | NCSP | WX723, WX780 | 2 |
| Psychotic disorders | ICD-10 | F20–F29 | 2, 3 |
| | ICPC-2 | P72 | 3 |
| Clozapine | ATC | N05AH02 | 4 |

ATC, Anatomical Therapeutic Chemical Classification System; ICD-10, International Statistical Classification of Diseases and Related Health Problems, tenth revision; ICPC-2, International Classification of Primary Care, second edition; NCSP, Nordic Nomesco Classification of Surgical Procedures.

Registers: 1, Special Reimbursement Register for Medicine Expenses; 2, Care Register for Health Care; 3, Register of Primary Health Care Visits; 4, Prescription Centre database.

**Supplementary Table 2:** Distribution of baseline characteristics and percentage vaccinated first with mRNA or adenovirus vector (AdV) vaccine, Finnish elderly aged 70+ years.

| | | Percentage vaccinated first with | |
|---|---|---|---|
| | Number of study subjects | mRNA vaccine | AdV vaccine |
| **Age in years** | | | |
| 70-74 | 353956 | 79 | 10 |
| 75-79 | 231915 | 86 | 4 |
| 80-89 | 258389 | 89 | 1 |
| 90+ | 56832 | 83 | 1 |
| **Sex** | | | |
| Male | 384170 | 83 | 6 |
| Female | 516922 | 85 | 5 |
| **Presence of medical conditions predisposing to severe Covid-19** | | | |
| No predisposing medical condition | 376668 | 83 | 6 |
| At least one highly predisposing medical condition | 314949 | 85 | 5 |
| At least one predisposing but no highly predisposing medical condition | 209475 | 85 | 5 |
| **In Helsinki-Uusimaa hospital district** | | | |
| No | 685004 | 83 | 6 |
| Yes | 216088 | 87 | 2 |
| **In long-term care** | | | |
| No | 851206 | 84 | 5 |
| Yes | 49886 | 85 | 0 |

**Supplementary Table 3:** Distribution of baseline characteristics and percentage vaccinated first with mRNA or adenovirus vector (AdV) vaccine, chronically ill aged 16–69 years.

| | | Percentage vaccinated first with | |
|---|---|---|---|
| | Number of study subjects | mRNA vaccine | AdV vaccine |
| **Age in years** | | | |
| 16-38 | 151887 | 33 | 4 |
| 39-51 | 151543 | 50 | 10 |
| 52-59 | 173280 | 58 | 19 |
| 60-64 | 138227 | 55 | 28 |
| 65-69 | 159589 | 32 | 54 |
| **Sex** | | | |
| Male | 403410 | 44 | 24 |
| Female | 371116 | 48 | 22 |
| **Presence of medical conditions predisposing to severe Covid-19** | | | |
| At least one highly predisposing medical condition | 329664 | 38 | 37 |
| At least one predisposing but no highly predisposing medical condition | 444862 | 51 | 12 |
| **In Helsinki-Uusimaa hospital district** | | | |
| No | 567944 | 46 | 22 |
| Yes | 206582 | 44 | 27 |

**Supplementary Table 4:** Crude and adjusted hazard ratios comparing the hazard of confirmed SARS-CoV-2 infection or Covid-19 hospitalization in study subjects who received exactly 1 or 2 doses of mRNA vaccine with the corresponding hazard in the unvaccinated, Finnish elderly aged 70+ years.

| | **SARS-CoV-2 infection** | | | | | | **Covid-19 hospitalization** | | | | | |
|---|---|---|---|---|---|---|---|---|---|---|---|---|
| | **Crude hazard ratio** | | | **Adjusted hazard ratio** | | | **Crude hazard ratio** | | | **Adjusted hazard ratio** | | |
| | Est. | LCI | UCI | Est. | LCI | UCI | Est. | LCI | UCI | Est. | LCI | UCI |
| **First dose** | | | | | | | | | | | | |
| 0-6 DSV | 0.773 | 0.624 | 0.956 | 0.669 | 0.541 | 0.827 | 0.465 | 0.270 | 0.801 | 0.425 | 0.246 | 0.733 |
| 7-13 DSV | 0.846 | 0.684 | 1.046 | 0.707 | 0.572 | 0.875 | 0.644 | 0.400 | 1.038 | 0.577 | 0.357 | 0.931 |
| 14-20 DSV | 0.766 | 0.607 | 0.966 | 0.618 | 0.490 | 0.780 | 0.831 | 0.538 | 1.285 | 0.734 | 0.473 | 1.138 |
| 21-27 DSV | 0.745 | 0.581 | 0.956 | 0.589 | 0.459 | 0.755 | 0.498 | 0.286 | 0.869 | 0.433 | 0.248 | 0.758 |
| 28-34 DSV | 0.693 | 0.527 | 0.911 | 0.534 | 0.406 | 0.701 | 0.470 | 0.263 | 0.841 | 0.414 | 0.231 | 0.742 |
| 35-41 DSV | 0.735 | 0.554 | 0.976 | 0.545 | 0.411 | 0.722 | 0.411 | 0.218 | 0.774 | 0.360 | 0.191 | 0.680 |
| 42+ DSV | 0.827 | 0.680 | 1.004 | 0.535 | 0.441 | 0.650 | 0.386 | 0.253 | 0.587 | 0.321 | 0.208 | 0.496 |
| **Second dose** | | | | | | | | | | | | |
| 0-6 DSV | 0.300 | 0.123 | 0.728 | 0.153 | 0.063 | 0.371 | 0.267 | 0.064 | 1.108 | 0.209 | 0.050 | 0.877 |
| 7+ DSV | 0.725 | 0.530 | 0.990 | 0.250 | 0.181 | 0.347 | 0.087 | 0.021 | 0.355 | 0.072 | 0.017 | 0.300 |

DSV, day since vaccination; Est., Point estimate; LCI, lower 95% confidence interval limit; UCI, upper 95% confidence interval limit

**Supplementary Table 5:** Crude and adjusted hazard ratios comparing the hazard of confirmed SARS-CoV-2 infection or Covid-19 hospitalization in study subjects who received exactly 1 or 2 doses of mRNA vaccine with the corresponding hazard in the unvaccinated, chronically ill aged 16–69 years.

| | **SARS-CoV-2 infection** | | | | | | **Covid-19 hospitalization** | | | | | |
|---|---|---|---|---|---|---|---|---|---|---|---|---|
| | **Crude hazard ratio** | | | **Adjusted hazard ratio** | | | **Crude hazard ratio** | | | **Adjusted hazard ratio** | | |
| | Est. | LCI | UCI | Est. | LCI | UCI | Est. | LCI | UCI | Est. | LCI | UCI |
| **First dose** | | | | | | | | | | | | |
| 0-6 DSV | 0.568 | 0.438 | 0.737 | 0.612 | 0.472 | 0.794 | 0.142 | 0.035 | 0.574 | 0.129 | 0.032 | 0.521 |
| 7-13 DSV | 0.605 | 0.462 | 0.792 | 0.664 | 0.506 | 0.870 | 0.242 | 0.077 | 0.763 | 0.218 | 0.069 | 0.687 |
| 14-20 DSV | 0.519 | 0.380 | 0.708 | 0.575 | 0.421 | 0.785 | 0.569 | 0.249 | 1.304 | 0.511 | 0.223 | 1.171 |
| 21-27 DSV | 0.532 | 0.378 | 0.749 | 0.587 | 0.417 | 0.827 | 0.120 | 0.017 | 0.862 | 0.106 | 0.015 | 0.764 |
| 28-34 DSV | 0.392 | 0.251 | 0.614 | 0.424 | 0.271 | 0.664 | 0.162 | 0.022 | 1.168 | 0.139 | 0.019 | 1.008 |
| 35-41 DSV | 0.386 | 0.222 | 0.670 | 0.411 | 0.236 | 0.714 | Not estimated | | | Not estimated | | |
| 42+ DSV | 0.686 | 0.506 | 0.928 | 0.857 | 0.632 | 1.161 | 0.411 | 0.131 | 1.287 | 0.410 | 0.131 | 1.288 |
| **Second dose** | | | | | | | | | | | | |
| 0-6 DSV | 0.115 | 0.016 | 0.817 | 0.127 | 0.018 | 0.906 | Not estimated | | | Not estimated | | |
| 7+ DSV | 0.217 | 0.139 | 0.336 | 0.227 | 0.146 | 0.353 | 0.095 | 0.013 | 0.678 | 0.100 | 0.014 | 0.710 |

DSV, day since vaccination; Est., Point estimate; LCI, lower 95% confidence interval limit; UCI, upper 95% confidence interval limit

**Supplementary Table 6:** Crude and adjusted hazard ratios comparing the hazard of confirmed SARS-CoV-2 infection or Covid-19 hospitalization in study subjects who received exactly 1 or 2 doses of adenovirus vector vaccine with the corresponding hazard in the unvaccinated, chronically ill aged 16–69 years.

| | **SARS-CoV-2 infection** | | | | | | **Covid-19 hospitalization** | | | | | |
|---|---|---|---|---|---|---|---|---|---|---|---|---|
| | **Crude hazard ratio** | | | **Adjusted hazard ratio** | | | **Crude hazard ratio** | | | **Adjusted hazard ratio** | | |
| | Est. | LCI | UCI | Est. | LCI | UCI | Est. | LCI | UCI | Est. | LCI | UCI |
| **First dose** | | | | | | | | | | | | |
| 0-6 DSV | 0.492 | 0.373 | 0.649 | 0.599 | 0.453 | 0.791 | 0.188 | 0.047 | 0.757 | 0.157 | 0.039 | 0.633 |
| 7-13 DSV | 0.656 | 0.514 | 0.836 | 0.804 | 0.630 | 1.027 | 1.163 | 0.666 | 2.029 | 0.961 | 0.548 | 1.685 |
| 14-20 DSV | 0.498 | 0.373 | 0.665 | 0.620 | 0.464 | 0.830 | 1.024 | 0.560 | 1.874 | 0.838 | 0.456 | 1.541 |
| 21-27 DSV | 0.605 | 0.458 | 0.799 | 0.764 | 0.578 | 1.011 | 0.414 | 0.154 | 1.112 | 0.335 | 0.124 | 0.903 |
| 28-34 DSV | 0.406 | 0.281 | 0.587 | 0.518 | 0.358 | 0.748 | 0.117 | 0.016 | 0.831 | 0.093 | 0.013 | 0.666 |
| 35-41 DSV | 0.505 | 0.350 | 0.730 | 0.635 | 0.438 | 0.919 | 0.815 | 0.360 | 1.843 | 0.638 | 0.281 | 1.449 |
| 42+ DSV | 0.401 | 0.318 | 0.507 | 0.497 | 0.393 | 0.630 | 0.564 | 0.327 | 0.973 | 0.423 | 0.243 | 0.736 |
| **Second dose** | | | | | | | | | | | | |
| 0-6 DSV | 0.687 | 0.220 | 2.147 | 1.375 | 0.439 | 4.309 | Not estimated | | | Not estimated | | |
| 7+ DSV | Not estimated | | | Not estimated | | | Not estimated | | | Not estimated | | |

DSV, days since vaccination; Est., Point estimate; LCI, lower 95% confidence interval limit; UCI, upper 95% confidence interval limit
