## Supplementary material for "Effectiveness of vaccination against SARS-CoV-2 infection and Covid-19 hospitalization among Finnish elderly and chronically ill – An interim analysis of a nationwide cohort study": Ethical concerns

To whom it may concern:

As director of the department for Health security of the Finnish Institute for Health and Welfare, I certify that:

- I am the competent auhority for assessing whether research requires institutionnal ethical review or if the Finnish communicable diseases law (Tartuntatautilaki 1227/2016)and the law on the duties of the Finnish institute for Health (Laki Terveyden ja hyvinvoinnin laitoksesta 668/2008) and Welfare allows the implementation of the research without seeking further ethical review.

- The research presented by Baum et al in "**Effectiveness of vaccination against SARS-CoV-2 infection and Covid-19 hospitalization among Finnish elderly and chronically ill – An interim analysis of a nationwide cohort study**" did not require further ethical review before implementation as its aim was to monitor vaccine effectiveness of infectious disease (Tartuntatautilaki 1227/2016).

Digitally signed by Mika Salminen
Date: 2021.06.22 09:27:11 +03'00'

Helsinki, June 17th 2021
Prof Mika Salminen
